# Adaptive Transfer Learning for Time-to-Event Modeling with Applications in Disease Risk Assessment

**DOI:** 10.1101/2025.01.14.25320536

**Authors:** Yuying Lu, Tian Gu, Rui Duan

## Abstract

To address the challenges for modeling time-to-event outcomes in small-sample settings, we propose a novel transfer learning approach, termed CoxTL, based on the widely used Cox proportional hazards model, accounting for potential covariate and concept shifts between source and target datasets. CoxTL utilizes a combination of density ratio weighting and importance weighting techniques to address multi-level data heterogeneity, including covariate and coefficient shifts between source and target datasets. Additionally, it accounts for potential model misspecification, ensuring robustness across a wide range of settings. We assess the performance of CoxTL through extensive simulation studies, considering data under various types of distributional shifts. Additionally, we apply CoxTL to predict End-Stage Renal Disease (ESRD) in the Hispanic population using electronic health record-derived features from the All of Us Research Program. Data from non-Hispanic White and non-Hispanic Black populations are leveraged as source cohorts. Model performance is evaluated using the C-index and Integrated Brier Score (IBS). In simulation studies, CoxTL demonstrates higher predictive accuracy, particularly in scenarios involving multi-level heterogeneity between target and source datasets. In other scenarios, CoxTL performs comparably to alternative methods specifically designed to address only a single type of distributional shift. For predicting the 2-year risk of ESRD in the Hispanic population, CoxTL achieves an increase in C-index up to 6.76% compared to the model trained exclusively on target data. Furthermore, it demonstrates up to 17.94% increase in the C-index compared to the state-of-the-art transfer learning method based on Cox model. The proposed method effectively utilizes source data to enhance time-to-event predictions in target populations with limited samples. Its ability to handle various sources and levels of data heterogeneity ensures robustness, making it particularly well-suited for real-world applications involving target populations with small sample sizes, where traditional Cox models often struggle.

## INTRODUCTION

Survival analysis is crucial across various disciplines, including medical research, finance, and social sciences, where modeling the time until an event of interest occurs provides actionable insights. The Cox Proportional Hazards (Cox) model (Cox, 1972) has emerged as a cornerstone in survival analysis due to its semi-parametric nature, allowing for flexible baseline hazard functions without imposing rigid assumptions. However, fitting a reliable Cox model becomes challenging in small-sample settings, where limited data can undermine coefficient estimation and reduce predictive accuracy. Such scenarios often arise in studies of rare diseases, subpopulations with limited representation, or specialized clinical trials.

Recent research has explored transfer learning as a promising solution to enhance model performance in small-sample scenarios. Transfer learning is a machine learning approach that leverages knowledge from one task (*the source*) to improve model performance in another related task with limited samples (*the target*) (Pan and Yang, 2009). Despite its widespread use in engineering (Niu et al., 2020), transfer learning has also been increasingly adopted in biomedical fields, including genetic risk prediction (Lu et al., 2024), disease risk score construction (Zhao et al., 2022; Tian et al., 2022), and drug discovery (Cai et al., 2020), where limited target data often pose significant challenges for model training. By leveraging information from a related source study, transfer learning enables more efficient learning and reduces data requirements. Techniques in transfer learning range from fine-tuning pre-trained models, domain adaptation strategies that adjust for differences between source and target datasets (Niu et al., 2020), to penalizing on the similarity between the target and the source parameter estimates (Li et al., 2016; Tian and Feng, 2023; Gu et al., 2022, 2023).

The application of transfer learning to survival analysis remains relatively underexplored. Limited studies have investigated how transfer learning can enhance survival models in time-to-event data. For example, Li et al. (2016) introduced *L*_2,1_-norm regularization to borrow strength from external data, while Wang et al. (2023) proposed a Kullback-Leibler-based Cox model to integrate individual-level time-to-event data with published risk scores. Similarly, Tran et al. (2022) employed cross-validation to selectively combine external and internal data, and Li et al. (2023) applied *L*_1_-norm penalized regression to handle heterogeneity in baseline hazards and coefficients. Although these methods address important aspects of inter-dataset variability, they often overlook covariate shifts—systematic differences in covariate distributions between source and target populations—which represent a common and significant source of data heterogeneity. Covariate shifts do not pose significant issues when the Cox model is correctly specified. However, in practice, the Cox model is often used as a “working model” to analyze the relationship between covariates and the hazard rate and to construct risk prediction models, even when its assumptions—such as proportional hazards—do not strictly hold (Sheu et al., 2023). Such misspecifications can amplify the effects of covariate shifts, introducing substantial biases that undermine the robustness and efficiency of transfer learning methods in survival analysis. Ignoring these shifts can, therefore, reduce model performance (Gerds and Schumacher, 2001). Recently, Fan et al. (2024) introduced a weighted Cox regression model to identify stable variables across cohorts for risk prediction, addressing distributional shifts in survival analysis. However, it requires a shared survival model across different cohorts, which may be hardly satisfied in real-world applications.

Related advancements in survival analysis highlight the growing importance of adapting models to diverse data settings. For instance, Ding et al. (2023) explored transfer learning in additive hazard models, while deep transfer learning approaches (Lao et al., 2017; Han et al., 2020; Lopez-Garcia et al., 2020) have demonstrated promise in capturing complex survival patterns. Additionally, methods linking primary cohorts with aggregate external data (Chen et al., 2024) underscore the potential of hybrid approaches to address data heterogeneity. Although distinct from our focus on covariate shift adjustments in the Cox framework, these developments reflect the broader landscape of innovation in survival analysis.

To address these limitations, we propose a novel transfer learning approach in the Cox model that accounts for multi-level data heterogeneity between source and target datasets, including covariate shifts, baseline hazard variation and coefficient heterogeneity. Our method estimates the density ratio between the target and source datasets and uses it to reweight the source data within the log-likelihood function of the Cox model for the target cohort. A tuning parameter is employed to adaptively control the influence of the source data during model fitting. By incorporating patient-level data from the source study, this two-step approach enables flexible calibration to address potential disparities in covariate, baseline hazard, and coefficients. Similar strategies of tuning parameters have been applied in other contexts, such as catalytic prior models (Huang et al., 2020; Li and Huang, 2023), where tuning parameters were used to regulate the impact of synthetic data. Through extensive simulation studies and a real-data application to predict the survival of ESRD in Hispanics, we demonstrate the effectiveness of our approach in improving robustness and predictive performance compared to existing methods, highlighting the value of addressing covariate shifts and other potential heterogeneity in data integration for survival models in diverse settings.

## METHODOLOGY

### Notations and problem setup

We consider a source cohort (denoted by 𝒮) with *N* ^𝒮^ independent observations and a target cohort (denoted by 𝒯) with *N*^*τ*^ independent observations. The target cohort is assumed to have a smaller sample size compared to the source cohort, such that *N* ^𝒮^ ≫ *N* ^*T*^. For the *i*-th observation in cohort *τ* ∈ {𝒮, 𝒯}, the covariates are represented as a *p*-dimensional vector 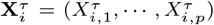, which is regarded as a *p* × 1 matrix, and the underlying survival time 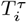 follows a proportional hazard model. The hazard function, which defines the instantaneous risk of the event occurring at time *t* given the covariates 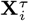, is expressed as

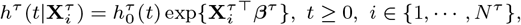

where 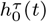 and ***β***^*τ*^ represent the baseline hazard function and the regression coefficient in cohort *τ*. For each sample, let 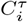 denote the censoring time, 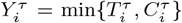 be the observed time, and 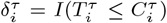 indicate whether the event of interest occurred before censoring. The observed data is 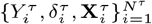, and we use 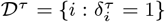 to denote the index set of case observations.

The regression parameter ***β***^*T*^ for the target cohort is traditionally estimated by maximizing the partial likelihood function

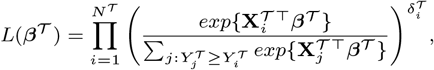

or equivalent, by maximizing the partial log-likelihood

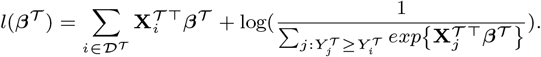

The solution 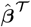 to the likelihood equation

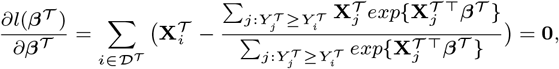

provides an estimator of 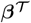. The baseline hazard function is estimated at the observed time 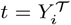 for the *i*-th observation by

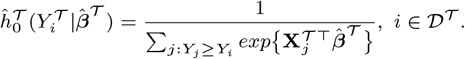

However, the accuracy of 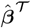 in this target-only model is limited by the small sample size of the target cohort. To address this limitation, we incorporate auxiliary observations 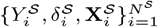 in a source cohort. These observations share certain similarities with the target data and, when used effectively, can enhance the Cox model’s performance by leveraging additional information. However, integrating source and target data is not straightforward due to potential heterogeneity between the cohorts. These include the potential difference in the distribution of **X** (referred to as covariate shift), baseline hazard functions, and regression coefficient. Without accounting for these disparities, naively pooling the two datasets can degrade model performance, leading to biased estimates or even negative transfers, where combining cohorts yields worse results than using the target data alone.

### Proposed two-step Cox transfer learning algorithm (CoxTL)

We propose a two-step transfer learning approach to leverage useful information from the source data while accounting for multi-level data heterogeneity, including the potential covariate shift and parameter disparity. A schematic diagram illustrating the workflow is shown in Figure 1.

**Fig. 1.**
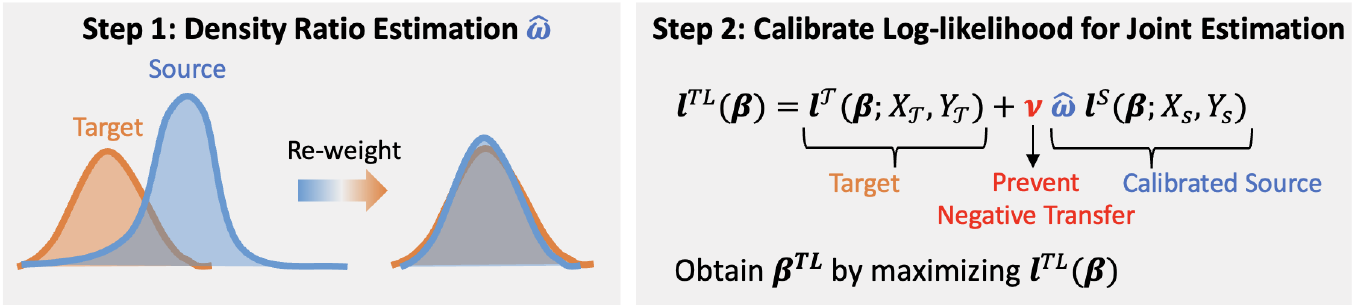
Diagram of the proposed 2-step transfer learning.

#### Step 1: Density ratio weighting to adjust for covariate shift

To adjust the potential covariate shift in the source data, we propose to re-weight the source data using the density ratio estimated between the target and the source, which has been considered in recent work (Lu et al., 2024; Li and Duan, 2024).

For cohort *τ* ∈ {𝒮, 𝒯}, let *P*^*τ*^ (·) denote the probability density function of **X** in cohort *τ*, and the density ratio model is defined as the probability density ratio between the target and the source populations, i.e.,

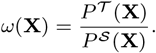

The function *ω*(**X**) can be estimated in flexibility using applicable data-adaptive methods, including exponential tilting models (Efron, 1978), generalized additive models (Hastie, 2017), and sieve methods (Grenander, 1981). Alternatively, each source can estimate its density function via methods such as wavelets density estimation (Donoho et al., 1996) or deep learning methods (Liu et al., 2021) and then obtain their ratios. In this paper, we illustrate the idea using the exponential tilting model. Denote **Z** = (1, **X**). We assume the density ratio model *ω*(**X**) follows the exponential tilting model with an underlying *p* + 1 dimensional parameter ***θ*** = (*θ*_0_, *θ*_1_, *· · ·*, *θ*_*p*_), i.e,.

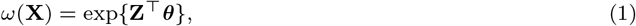

where *θ*_0_ is a normalizing constant or can be considered as an “intercept”. When *ω*(**X**) is correctly specified, the relationship 𝔼 ^𝒯^ [*f* (**X**)] = 𝔼 ^𝒮^ [*ω*(**X**) *· f* (**X**)] holds for any function *f* (**X**), where 𝔼 ^*τ*^ [*·*] is the expectation with respect to *P*^*τ*^ (·), *τ ∈* {𝒮, 𝒯}. By inserting the assumed exponential tilting model from equation (1), we have the following equation hold:

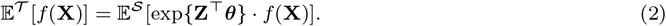

By letting *f* (**X**) = **Z**, we have ***θ*** = arg min_***θ***_ {𝔼 ^𝒮^ [exp **Z**^*⊤*^***θ***] *−*𝔼 ^𝒯^ [**Z**^*⊤*^***θ***]}, which implies ***θ*** is the root of the first derivative of the objective function. Therefore, the minimizer ***θ*** satisfies equation (2). In practice, this can be obtained through

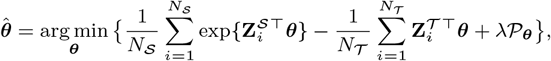

where 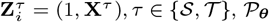 can be any penalty function encouraging certain structures on ***θ***, and *λ* is the tuning parameter. In this study, we use L2 regularization as the penalty, i.e., 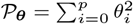 in simulation and real data analysis.

#### Step 2: Calibrate log-likelihood to leverage source data while mitigating potential bias

Given the density ratio model,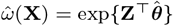, estimated in Step 1, the source data can be re-weighted to construct a calibrated log-likelihood function that incorporates source information while addressing covariate disparity. Additionally, we introduce a scalar hyperparameter to control the overall heterogeneity, accounting for potential differences in both the baseline hazard and the regression parameters.

Specifically, we propose a re-weighted partial log-likelihood function via transfer learning, *l*^TL^(***β***), consisting of two parts

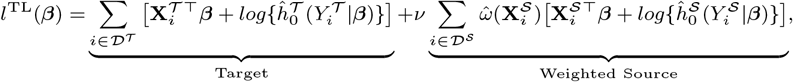

where

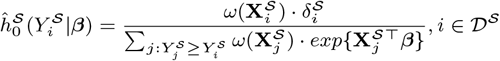

is the calibrated source baseline hazard estimation. The hyperparameter *ν* is designed to control the degree of information sharing from the source cohort to the target cohort, thereby preventing negative transfer learning caused by significant heterogeneity between the target and the source data, or potential model misspecifications. The value of *ν* can be selected using cross-validation.

Finally, the proposed transfer learning estimator of ***β***^*T*^ and the corresponding cumulative baseline hazard function can be obtained through

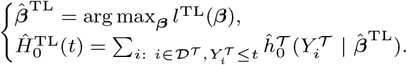

## SIMULATION

### Simulation settings

We evaluate the performance of CoxTL under scenarios with and without covariate shifts, incorporating varying levels of model heterogeneity, as well as altering the baseline hazard function. We begin with generating p-dimensional covariates **X** = (*X*_1_, *· · ·, X*_*p*_), for a large number of samples size *N* to create a covariate pool. A binary source indicator, *R*, is then created to denote whether the data belongs to the source population. Based on *R*, we divide the samples into cohort 𝒮 and 𝒯. For each cohort, the survival outcomes are generated using the covariates, the respective hazard function, and censoring time. Specifically, the disease status *δ* and the event time *Y* are determined based on the hazard function and censoring time. Finally, we randomly select *N*^*τ*^ samples from cohort *τ* as our final dataset, *τ* ∈ {𝒮, 𝒯} and independently choose *N* ^test^ target samples as the test dataset. The initial data sample size *N* is set to be large enough to ensure there are sufficient samples in each cohort to select.

More specifically, we first generate a large dataset of size *N* with corresponding *p* dimensional covariates **X** = (*X*_1_, *· · ·*, *X*_*p*_), where

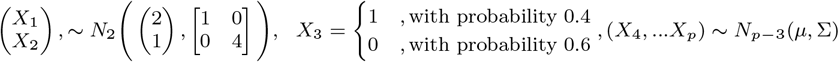

where *µ* = (0.5, 1, 0.5, 1, ….), Σ_*ij*_ = 0.1^|*i−j*|^, and *i, j ∈* {1, *· · ·*, *p*}. We vary dimension *p ∈* {10, 30, 50}.

To control the existence of covariate shift, we use the logistic regression model

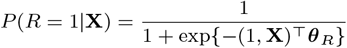

to generate *R*, where ***θ***_*R*_ = (*θ*_*R*0_, *· · ·*, *θ*_*Rp*_) is a (*p* + 1)-dimensional parameter. We can compute the density ratio through the Bayesian formula

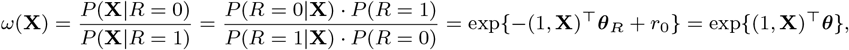

where 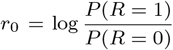 is a constant. Therefore, the density ratio *ω*(**X**) follows the suggested exponential tilting model with the true parameter ***θ*** = (*− θ*_*R*0_ +*r*_0_, *− θ*_*R*1_, *· · ·*,*−θ*_*Rp*_). We consider the following two logistic models with different ***θ***_*R*_:

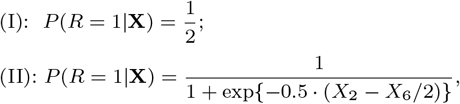

where (I) leads to no covariate shift while (II) introduces a covariate shift between source and target cohorts. For the samples in the target cohort, we impute their survival time T with the hazard function

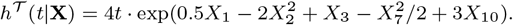

To mimic the potential heterogeneous in the baseline hazard model, we consider four hazard functions for the source cohort:

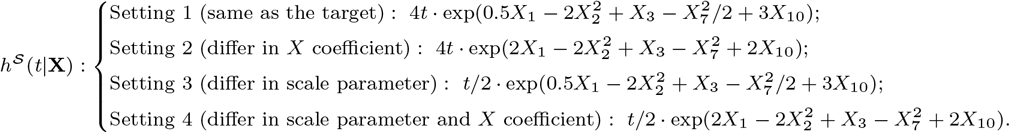

The censoring time *C* for both cohorts follows Unif(0, 3.55). Finally, we get the status *δ* = *I* {*T ≤C*} and event time *Y* = min {*T, C*} for each sample.

We set the initial data size *N* = 20000, the target sample size *N* ^𝒯^ = 200, the source sample size *N* ^𝒮^ = 4000 and the test set size *N* ^test^ = 500. For each simulation scenario, we train all the models using the same training dataset and test the model performance on the test data. We set the parameter *λ* = 1 as the default value for the *L*_2_ penalty *𝒫*_***θ***_ in step 1, and the hyperparameter *ν* in step 2 of CoxTL is selected through 5-fold cross-validation.

We compare the performance of the proposed CoxTL with the following five methods that are applicable in the same setting: (1) **Cox Orac**: Cox model fitted using a large number of target samples of size *N* ^orac^ = 5000 following the true data generating mechanism; (2) **Cox T**: a classical Cox model using only the limited target sample of size *N* ^𝒯^; (3) **Cox S**: Cox model using the source sample with a large number of samples of size *N* ^𝒮^;(4) **Cox Str**: stratified Cox model using the target samples of size *N* ^𝒯^ and the source samples of size *N* ^𝒮^; and (5) **TransCox**: an existing transfer learning approach to leverage source information through penalizing the difference in both the baseline hazard and the parameters between the target and the source data (Li et al., 2023).

In each setting, we replicate 100 simulations and calculate the C-index and the Integrated Brior Score (IBS)(Graf et al., 1999) to evaluate the model performance on the test set each time. The IBS is defined as 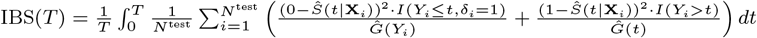, where *Ĝ*(*t*) = *P* (*C > t*) is the estimator of the conditional survival function of the censoring times calculated using the Kaplan-Meier method and C denotes the censoring time.

### Simulation results

The simulation results show that CoxTL demonstrates its strength in calibrating potential model heterogeneity between the two populations. It significantly outperforms other methods, particularly when the source hazard function differs substantially from that of the target cohort (as seen in Settings 2 and 4). Moreover, CoxTL exhibits a distinct advantage when the covariate shift exists between the cohorts, a scenario that closely reflects real-world situations. The results are shown in Tables 1 - 2 and Figures 2 - 5.

**Table 1.** Average values of C-index and IBS for Cox_Orac, Cox_T, Cox_S,Cox_Str, TransCox and CoxTL over 100 simulations under four settings without covariate shift by varying dimension p *∈* {10, 30, 50}. The best result in each setting is highlighted in bold.

|  | C-index | IBS | C-index | IBS | C-index | IBS | C-index | IBS |
| --- | --- | --- | --- | --- | --- | --- | --- | --- |
| $p = 10$ | Setting 1 | | Setting 2 | | Setting 3 | | Setting 4 | |
| Cox_Orac | 0.7933 | 0.1541 | 0.7926 | 0.1542 | 0.7916 | 0.1553 | 0.7980 | 0.1548 |
| Cox_T | 0.7658 | 0.1631 | 0.7654 | 0.1634 | 0.7642 | 0.1646 | 0.7734 | 0.1632 |
| Cox_S | 0.7934 | **0.1541** | 0.7522 | 0.1720 | 0.7857 | 0.1764 | 0.7504 | 0.1686 |
| Cox_Str | **0.7936** | 0.1555 | 0.7558 | 0.1665 | **0.7860** | **0.1564** | 0.7545 | 0.1678 |
| TransCox | 0.7767 | 0.1596 | 0.7714 | **0.1609** | 0.7725 | 0.1613 | 0.7750 | **0.1620** |
| Cox_TL | 0.7928 | 0.1556 | **0.7749** | 0.1610 | 0.7852 | 0.1568 | **0.7769** | 0.1622 |
| $p = 30$ | Setting 1 | | Setting 2 | | Setting 3 | | Setting 4 | |
| Cox_Orac | 0.7910 | 0.1555 | 0.7938 | 0.1552 | 0.7923 | 0.1557 | 0.7907 | 0.1534 |
| Cox_T | 0.7200 | 0.1875 | 0.7243 | 0.1865 | 0.7222 | 0.1875 | 0.7265 | 0.1844 |
| Cox_S | 0.7896 | **0.1558** | 0.7515 | 0.1724 | 0.7846 | 0.1783 | 0.7443 | 0.1672 |
| Cox_Str | **0.7900** | 0.1577 | 0.7550 | 0.1680 | **0.7849** | **0.1580** | 0.7491 | 0.1659 |
| TransCox | 0.7357 | 0.1769 | 0.7344 | 0.1758 | 0.7387 | 0.1764 | 0.7355 | 0.1741 |
| Cox_TL | 0.7881 | 0.1578 | **0.7630** | **0.1654** | 0.7810 | 0.1588 | **0.7591** | **0.1635** |
| $p = 50$ | Setting 1 | | Setting 2 | | Setting 3 | | Setting 4 | |
| Cox_Orac | 0.7885 | 0.1557 | 0.7894 | 0.1531 | 0.7858 | 0.1558 | 0.7856 | 0.1546 |
| Cox_T | 0.6905 | 0.2159 | 0.6914 | 0.2123 | 0.6926 | 0.2168 | 0.6954 | 0.2143 |
| Cox_S | 0.7848 | **0.1562** | 0.7467 | 0.1731 | 0.7759 | 0.1761 | 0.7410 | 0.1681 |
| Cox_Str | **0.7850** | 0.1577 | 0.7504 | 0.1663 | **0.7766** | **0.1588** | 0.7455 | 0.1670 |
| TransCox | 0.7139 | 0.1916 | 0.7083 | 0.1871 | 0.7235 | 0.1868 | 0.7104 | 0.1903 |
| Cox_TL | 0.7821 | 0.1586 | **0.7552** | **0.1651** | 0.7740 | 0.1597 | **0.7511** | **0.1661** |

**Fig. 2.**
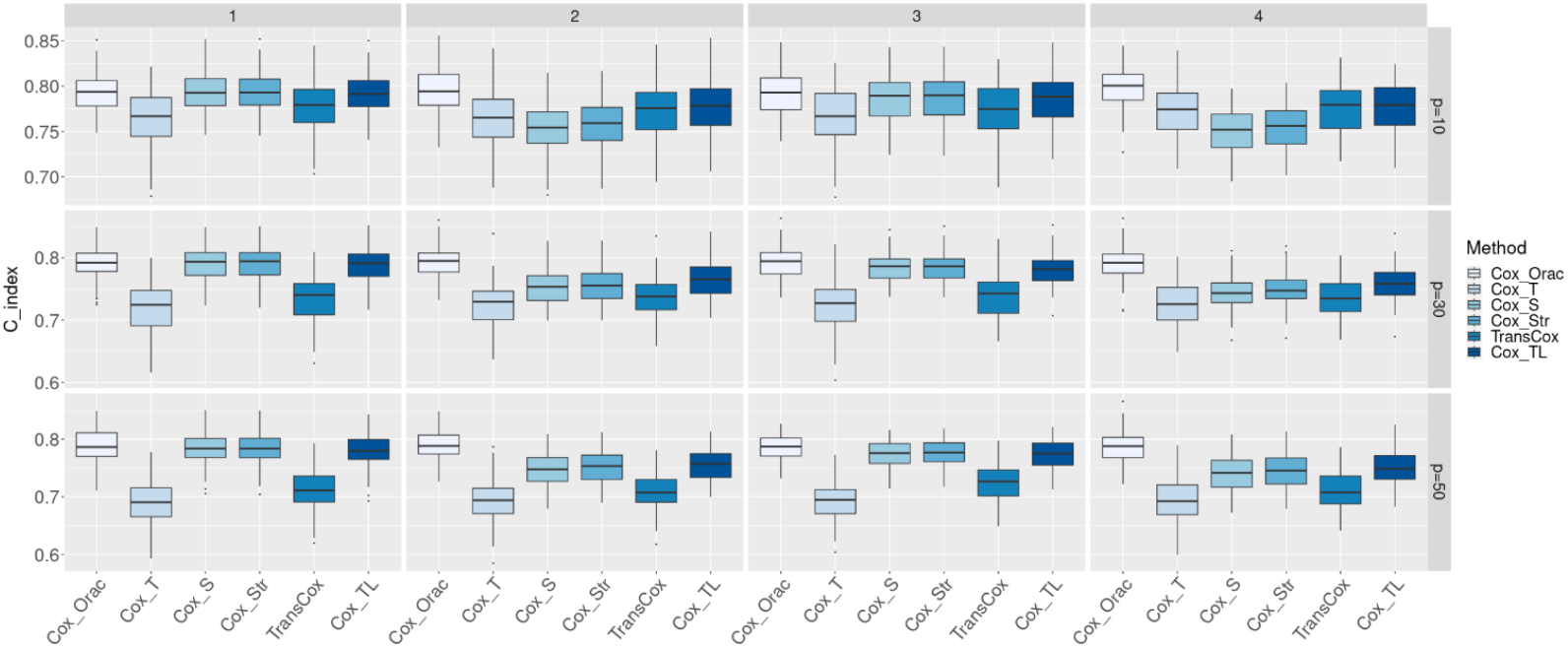
Boxplot of the C-index over 100 simulations under the setting without covariate shift between the target and the source.

Specifically, we first look at results regarding data without covariate shifts. As shown in Figure 2, CoxTL consistently achieves high C-index values across all settings. Specifically, its performance is close to or slightly below Cox Orac, which represents the oracle model assuming perfect knowledge of the underlying data-generating process. The advantage of CoxTL becomes more apparent in Settings 2 and 4, where the regression model difference between the source and target cohorts is more pronounced. This demonstrates the robustness of CoxTL in effectively leveraging source data while maintaining predictive accuracy. Additionally, while the performance of Cox S, Cos Str and TransCox varies with the value of the covariate dimension *p*, CoxTL has consistently satisfying accuracy whenever the dimension *p* is large or small.

Figure 3 highlights that CoxTL achieves nearly the lowest IBS in most scenarios, indicating its superior calibration compared to other methods. Notably, CoxTL outperforms TransCox and Cox Str in Settings 2 and 4, where heterogeneity exists in either the baseline hazard functions or the regression parameters between cohorts. In setting 3, where only baseline hazard function differs between cohorts, CoxTL has comparable performance with Cox Str and still overwhelms TransCox. Besides, CoxTL illustrates consistently great IBS with different covariate dimensions *p*. These underscore the model’s ability to balance robustness and efficiency in the absence of covariate shifts.

**Fig. 3.**
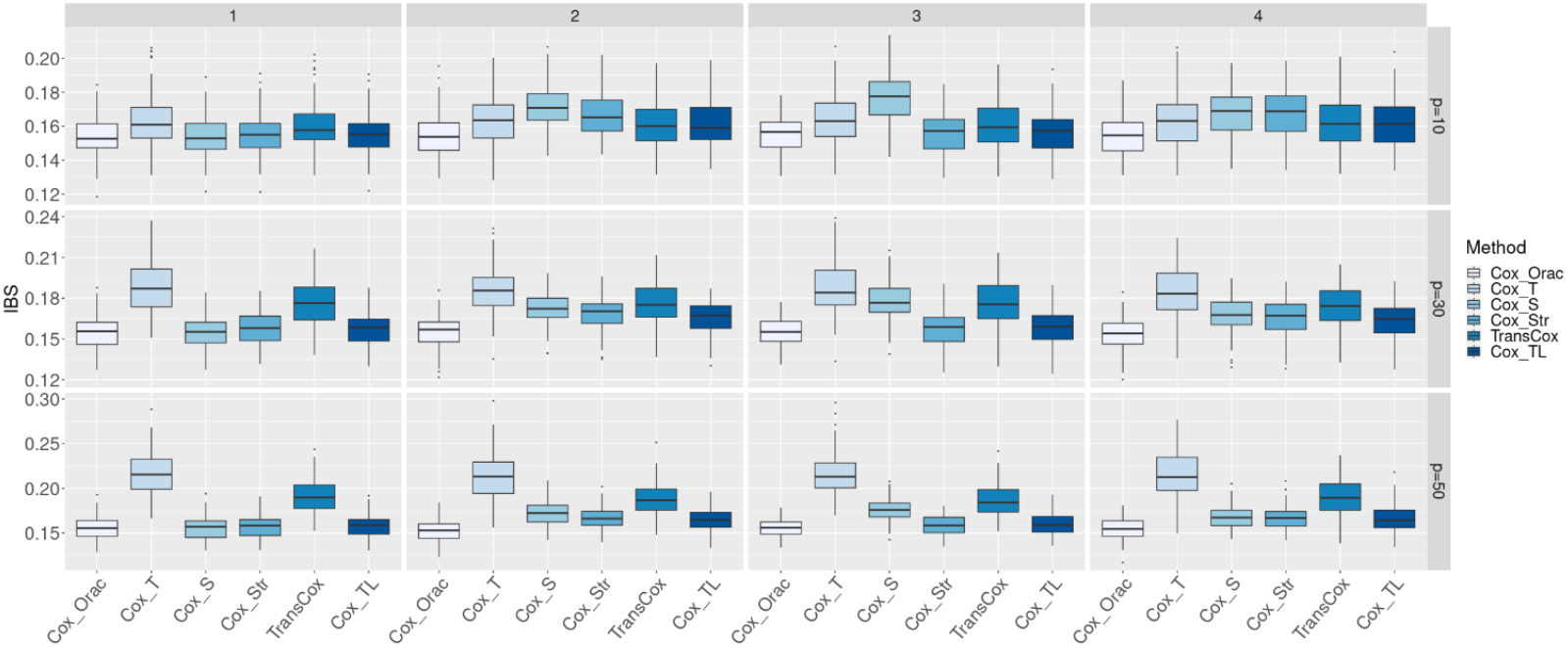
Boxplot of the IBS over 100 simulations under the setting without covariate shift between the target and the source.

Table 1 provides a quantitative summary of the C-index and IBS values. CoxTL consistently achieves results close to those of Cox Orac, particularly in high-dimensional settings (*p*=30 and *p*=50). This further validates the utility of CoxTL in accurately modeling survival outcomes without covariate shift.

Next, we look at the setting where covariate shifts exist between the target and the source. As shown in Figure 4, CoxTL significantly outperforms other methods under covariate shift scenarios. Its performance remains robust across all settings, with a marked advantage in Settings 2 and 4, where covariate distributions and regression parameters differ significantly between source and target cohorts. This highlights the model’s ability to adjust for covariate shifts while maintaining high predictive accuracy.

**Fig. 4.**
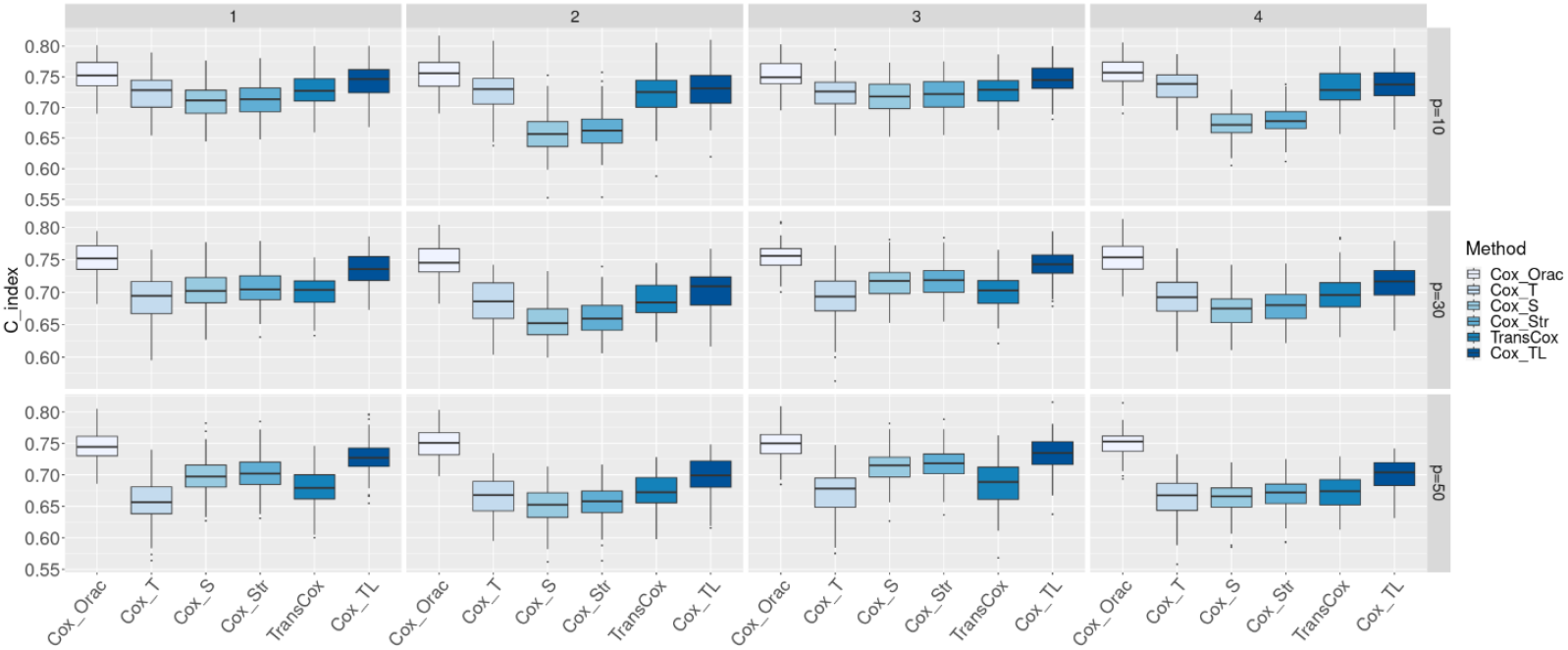
Boxplot of the C-index over 100 simulations under the setting with covariate shift between the target and the source.

Figure 5 demonstrates that CoxTL achieves superior calibration compared to other methods under covariate shift. Notably, the model maintains lower IBS values across all settings, with a particularly pronounced advantage in Settings 3 and 4. This indicates that CoxTL effectively addresses the challenges of covariate shifts in survival analysis.

**Fig. 5.**
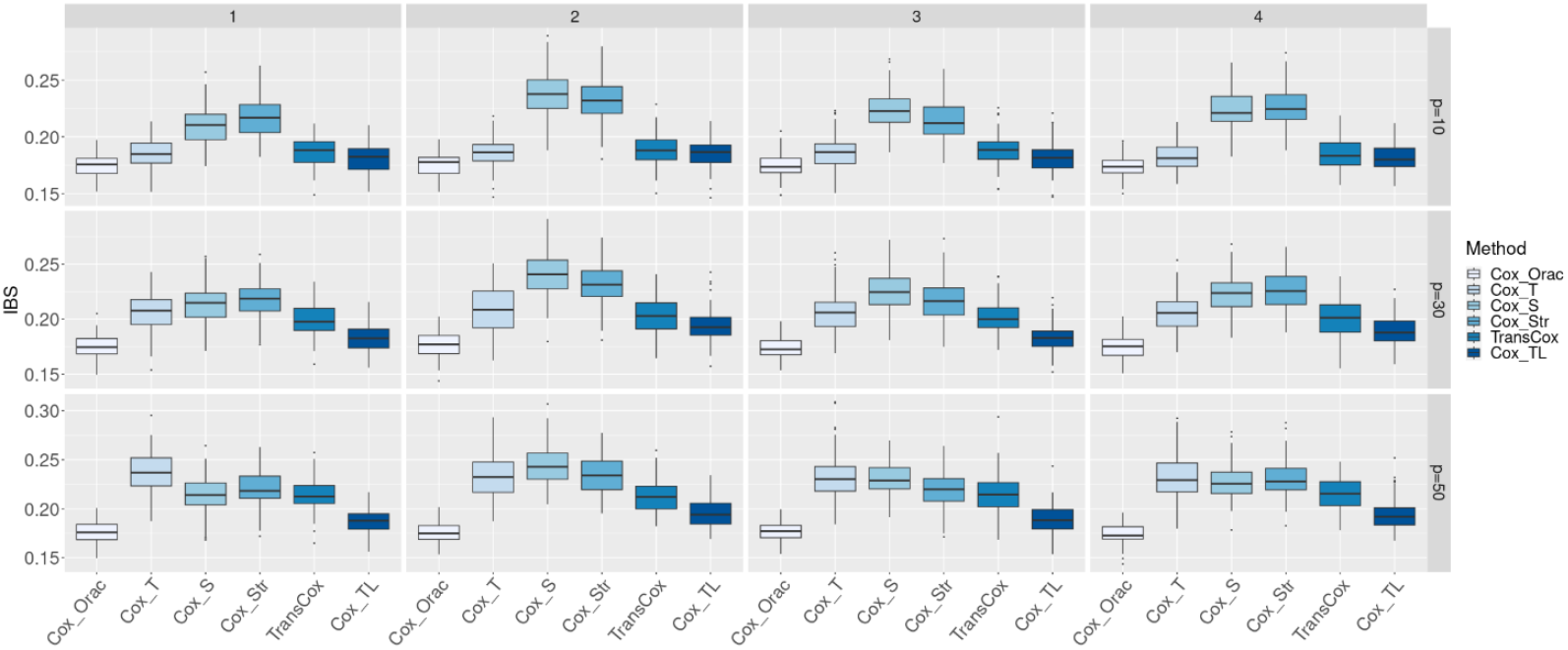
Boxplot of the IBS over 100 simulations under the setting with covariate shift between the target and the source.

Table 2 corroborates the findings from Figures 4 and 5. CoxTL consistently delivers the best performance in terms of both C-index and IBS, particularly in high-dimensional settings (*p*=30 and *p*=50). Its ability to outperform TransCox and Cox Str in these scenarios highlights its effectiveness in handling covariate shifts and leveraging source data for robust risk prediction.

**Table 2.** Average values of C-index and IBS for Cox_Orac, Cox_T, Cox_S, Cox_Str, TransCox and CoxTL over 100 simulations under four settings with covariate shift by varying dimension p *∈* {10, 30, 50}. The best result in each setting is highlighted in bold.

|  | C-index | IBS | C-index | IBS | C-index | IBS | C-index | IBS |
| --- | --- | --- | --- | --- | --- | --- | --- | --- |
| $p = 10$ | Setting 1 | | Setting 2 | | Setting 3 | | Setting 4 | |
| Cox_Orac | 0.7533 | 0.1745 | 0.7555 | 0.1748 | 0.7529 | 0.1749 | 0.7582 | 0.1736 |
| Cox_T | 0.7263 | 0.1850 | 0.7274 | 0.1853 | 0.7253 | 0.1861 | 0.7358 | 0.1819 |
| Cox_S | 0.7092 | 0.2096 | 0.6597 | 0.2370 | 0.7185 | 0.2238 | 0.6735 | 0.2228 |
| Cox_Str | 0.7123 | 0.2161 | 0.6652 | 0.2317 | 0.7217 | 0.2141 | 0.6795 | 0.2266 |
| TransCox | 0.7284 | 0.1860 | 0.7225 | 0.1885 | 0.7268 | 0.1886 | 0.7315 | 0.1848 |
| Cox_TL | **0.7436** | **0.1809** | **0.7285** | **0.1850** | **0.7448** | **0.1817** | **0.7381** | **0.1814** |
| $p = 30$ | Setting 1 | | Setting 2 | | Setting 3 | | Setting 4 | |
| Cox_Orac | 0.7507 | 0.1744 | 0.7468 | 0.1768 | 0.7543 | 0.1742 | 0.7540 | 0.1744 |
| Cox_T | 0.6912 | 0.2066 | 0.6852 | 0.2097 | 0.6928 | 0.2062 | 0.6928 | 0.2053 |
| Cox_S | 0.7032 | 0.2135 | 0.6541 | 0.2412 | 0.7149 | 0.2258 | 0.6724 | 0.2232 |
| Cox_Str | 0.7067 | 0.2174 | 0.6599 | 0.2324 | 0.7181 | 0.2165 | 0.6782 | 0.2263 |
| TransCox | 0.6999 | 0.1997 | 0.6886 | 0.2024 | 0.7007 | 0.2016 | 0.6978 | 0.2001 |
| Cox_TL | **0.7350** | **0.1833** | **0.7040** | **0.1938** | **0.7422** | **0.1829** | **0.7148** | **0.1891** |
| $p = 50$ | Setting 1 | | Setting 2 | | Setting 3 | | Setting 4 | |
| Cox_Orac | 0.7464 | 0.1761 | 0.7501 | 0.1752 | 0.7481 | 0.1772 | 0.7493 | 0.1740 |
| Cox_T | 0.6583 | 0.2367 | 0.6669 | 0.2322 | 0.6702 | 0.2322 | 0.6630 | 0.2326 |
| Cox_S | 0.6994 | 0.2144 | 0.6520 | 0.2449 | 0.7128 | 0.2303 | 0.6630 | 0.2268 |
| Cox_Str | 0.7031 | 0.2208 | 0.6574 | 0.2348 | 0.7165 | 0.2198 | 0.6689 | 0.2302 |
| TransCox | 0.6786 | 0.2148 | 0.6736 | 0.2130 | 0.6862 | 0.2154 | 0.6715 | 0.2157 |
| Cox_TL | **0.7279** | **0.1874** | **0.6986** | **0.1958** | **0.7331** | **0.1890** | **0.7017** | **0.1946** |

## REAL DATA APPLICATION

Chronic kidney disease (CKD) poses significant challenges due to its heterogeneous progression, varied clinical presentations, and the competing risk of cardiovascular mortality (Webster et al., 2017). While CKD progression is often gradual and can be modulated by interventions, early identification of high-risk patients remains essential to improve overall outcomes, such as reducing cardiovascular mortality and improving the management of ESRD. This is particularly important in populations with limited access to healthcare, where delays in diagnosis and treatment could accelerate progression to ESRD.

Traditional End-Stage Renal Disease (ESRD) risk prediction tools, such as the Kidney Failure Risk Equation (Peeters et al., 2013), rely heavily on laboratory-based metrics like estimated glomerular filtration rate (eGFR). However, these methods often neglect the potential advantages of including additional contextual information, such as diagnosis codes from electronic health records (EHR). Furthermore, risk prediction models often fail to generalize well across Hispanic subgroups due to differences in genetic, socioeconomic, and healthcare access factors (García and Ailshire, 2019; Maldonado et al., 2023; Errisuriz et al., 2024). The limited representation of Hispanic individuals in large prospective CKD studies further exacerbates these disparities (Lora et al., 2009), making it crucial to develop transfer learning approaches that leverage information from better-represented populationsMoreover, due to the small sample size of Hispanics with ESRD in many datasets, model estimates often suffer from low statistical power, limiting their reliability in clinical decision-making (Lora et al., 2009). To address these issues, we apply our proposed CoxTL framework to improve ESRD risk prediction for Hispanics. Our approach integrates EHR-derived features with eGFR-based filtering, leveraging transfer learning to calibrate data from non-Hispanic Black and White cohorts to mitigate data scarcity and heterogeneity.

We demonstrate the method using the Controlled Tier Dataset from the All of Us Research Program (version 7, C2022Q4R11, released December 5, 2023). This dataset represents a broader and more diverse population, including individuals who may not have access to specialized nephrology care. The dataset contains data collected through July 1, 2022 and initially includes 127,783 participants, of whom 1,655 are identified as having ESRD. This group consists of 51 Hispanics, 788 Non-Hispanic Blacks, and 762 Non-Hispanic Whites. The remaining 126,128 participants without ESRD include 2,990 Hispanics, 28,659 Non-Hispanic Blacks, and 90,514 Non-Hispanic Whites. We use Hispanics as the target population and consider two source cohorts: non-Hispanic Black, and non-Hispanic White.

To focus on individuals at higher risk of ESRD, we include only those with impaired kidney function, defined as having an eGFR below 65, calculated using the Modification of Diet in Renal Disease equation (Levey et al., 2009). After filtering, the dataset contains 15,566 participants: 203 Hispanics, 2,619 non-Hispanic Blacks, and 12,504 non-Hispanic Whites. Among these, ESRD measurements are available for 33 Hispanics, 408 non-Hispanic Blacks, and 319 non-Hispanic Whites.

Data are right-censored at 2 years and 5 years to evaluate short-term and long-term ESRD risks within these time frames. The All of Us dataset does not follow a fixed visit schedule as seen in prospective cohort studies. In our dataset, participants had a median of 6.31 (IQR: 2.89–10.88) recorded follow-up years between baseline and ESRD occurrence or censoring. The time gaps between follow-ups varied by individual, with a summary of follow-up time distribution presented in Supplementary Figure 2.

We include a comprehensive set of clinical variables available in the All of Us dataset, including demographic factors (age, sex, race/ethnicity), laboratory biomarkers (serum creatinine, eGFR, hemoglobin A1c), comorbid conditions (hypertension, diabetes, cardiovascular disease), and prescribed medications (angiotensin-converting enzyme inhibitors, angiotensin II receptor blockers, diuretics, and sodium-glucose cotransporter-2 inhibitors). These covariates were selected based on their known relevance in CKD progression and ESRD risk prediction. Full details of the covariate list are provided in Supplementary Table 1. The index date for each participant is defined as the date of their first serum creatinine measurement. The initial data set includes potential predictors and clinical diagnosis code counts measured in a one-year retrospective feature window prior to the index date. The event time is defined as the first diagnosis record of ESRD, and participants diagnosed with ESRD before the index date are excluded. Count data are normalized using log(*x* + 1) transformation followed by scaling. To mitigate multicollinearity, variables contributing to redundancy are removed, resulting in 60 predictors included in the final model training.

We randomly select 60% of the target samples as training data for model training while keeping the remaining 40% target samples for model testing, and we repeat this random data splitting 30 times for a robust performance evaluation. Each time, we use target training data and all source samples to estimate the density ratio and construct the re-weighted log-likelihood for model fitting. We set the parameter *λ* = 1 as the default value for the *L*_2_ penalty *𝒫* _***θ***_ in step 1 and the hyperparameter *ν* in step 2 of CoxTL is selected through 3-fold cross-validation. The C-index and IBS are calculated to assess the model performance when applied to the testing data. Meanwhile, we also fit Cox T, Cox Str, Cox S, and TransCox models, and compare their C-index and IBS with our proposed method.

Figure 6 illustrates the original and weighted distribution of covariates–BMI, age, body weight, and height– for the source (White) and target cohorts based on a single random data splitting. The results demonstrate the effectiveness of the estimated density ratio model to address the covariate shift between the source and target cohorts. Specifically, the weighted distributions show reduced differences in mean covariate values, demonstrating that the density ratio model successfully adjusts for heterogeneity in covariates.

**Fig. 6.**
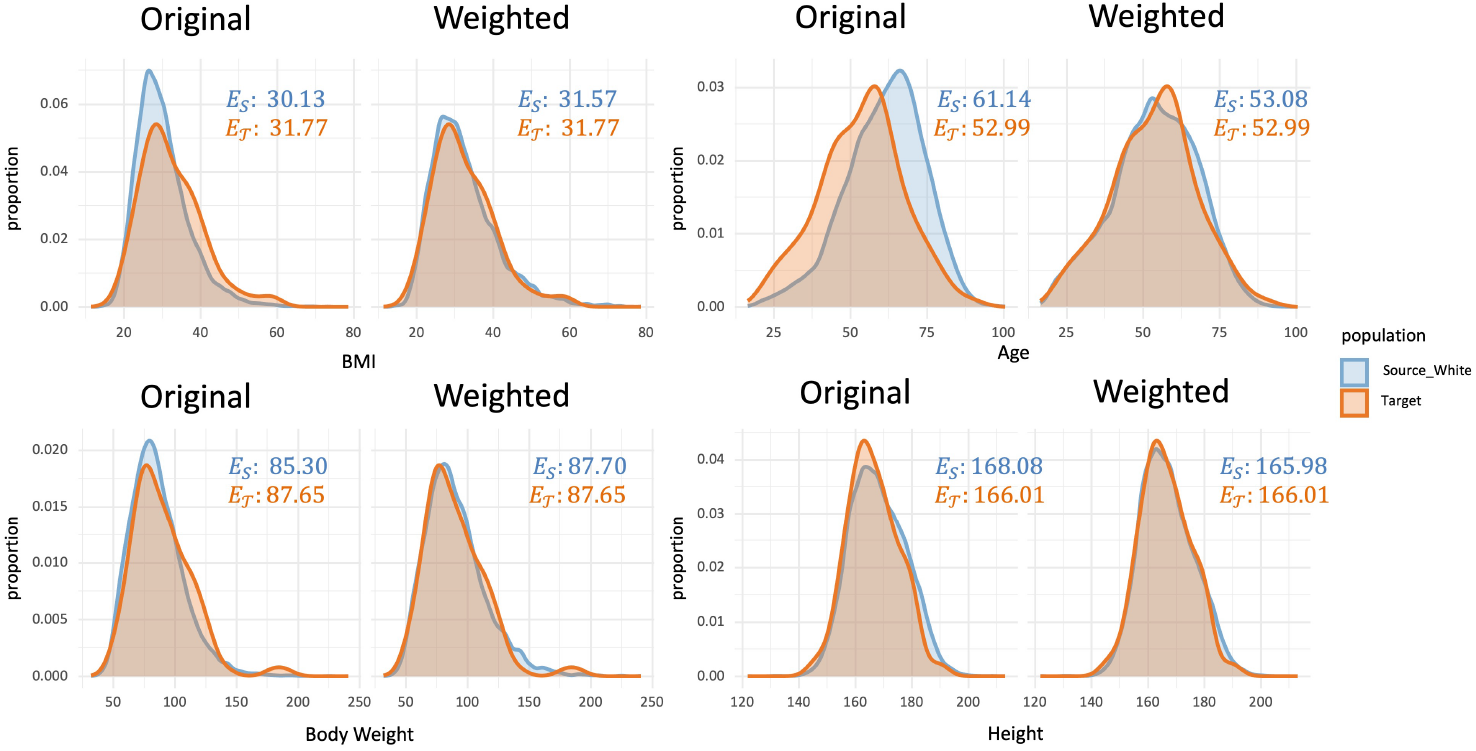
Comparison of the original and weighted distribution of BMI, Age, Body Weight, and Height, between the source (White) and target population from one random data splitting. We also show the mean value of each covariate in the source (*E*_*S*_) and in target (*E*_*T*_) populations.

Table 3 summarizes the predictive performance of CoxTL compared to four alternative methods for predicting 2-year and 5-year ESRD risks in Hispanics, evaluated using average C-index and IBS. The results demonstrate that CoxTL consistently outperforms all comparison methods across both short-term (2-year) and long-term (5-year) predictions. For example, when leveraging Black source data, CoxTL achieves a 2-year C-index of 0.8486 and IBS of 0.1853, marking significant improvements over the next-best method, Cox T, with a C-index of 0.7949 and IBS of 0.1642. Similarly, when using White source data, CoxTL achieves a C-index of 0.7782 and IBS of 0.1073, outperforming other methods in the same setting. Notably, CoxTL performs particularly well in short-term prediction (2-year), where it achieves both higher C-index values and relatively low IBS compared to competing methods. For long-term predictions (5-year), CoxTL continues to demonstrate robust performance, achieving the best C-index in nearly all settings, reflecting its ability to handle the heterogeneity between source and target cohorts effectively.

**Table 3.**
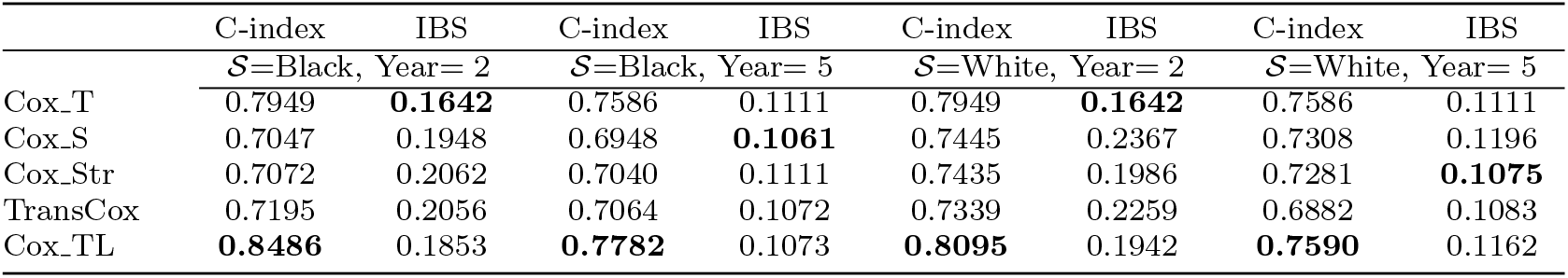
(Single-source CoxTL) Average values of C-index and IBS for Cox_T, Cox_S, Cox_Str, TransCox, and CoxTL over 30 replicates for 2-year and 5-year risk prediction, respectively. We present the results of single-source transfer learning using non-Hispanic blacks (Black) and non-Hispanic whites (White) as the source, respectively. The best result in each setting is highlighted in bold.

To further enhance the model’s applicability, we extend CoxTL to a multi-source framework, leveraging data from both Black and White populations simultaneously. In this multi-source framework, separate density ratio models are fit for each source population, and their weighted log-likelihoods are jointly incorporated into the proposed log-likelihood function *l*^TL^, for estimating ***β***^TL^ (See Section 1 ‘Multi-Source Framework’ in Supplementary Material for the explicit form of the log-likelihood function). Table 4 shows the performance of the multi-source CoxTL (denoted as CoxTL All), comparing with target-only Cox method (Cox T) and two single-source CoxTL methods using Black (CoxTL Black) or White (CoxTL White) population as the source population. CoxTL All achieves a higher AUC than Cox T, showing its capability to improve prediction in Hispanics. While CoxTL All does not consistently outperform the best single-source method in all settings, it shows greater robustness compared to the single-source approaches, particularly in balancing performance across diverse source populations. Notably, when single-source CoxTL methods don’t show significant performance compared with the target-only Cox method, for example, the 5-year prediction, multi-source CoxTL has the potential to achieve higher risk prediction accuracy.

**Table 4.**
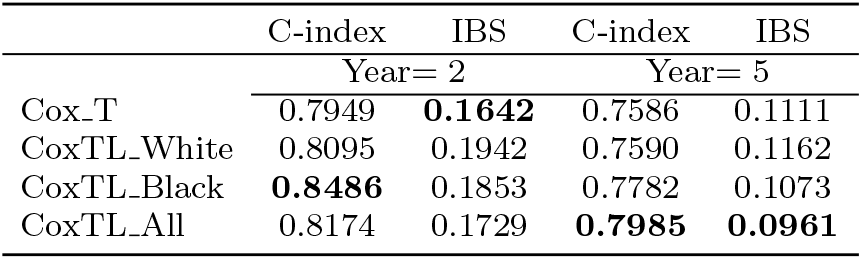
(Multi-source CoxTL) Average values of C-index and IBS for Cox_T, CoxTL_White, CoxTL_Black, and CoxTL_All over 30 replicates for 2-year and 5-year risk prediction, respectively. We present the results of single-source transfer learning using non-Hispanic blacks (CoxTL Black) and non-Hispanic whites (CoxTL_White) as the source, respectively. In addition, we present the results of multi-source CoxTL (CoxTL_All) that transfers information from both non-Hispanic blacks and non-Hispanic whites to Hispanics. The best result in each setting is highlighted in bold.

## DISCUSSION

This study introduces a novel Cox transfer learning approach, named CoxTL, that enhances prediction accuracy for the Cox model in a target cohort by effectively leveraging information from a related source study, through the adjustment of covariate shifts. Furthermore, our method introduces a tuning parameter to regulate the source likelihood’s influence, providing flexibility and control over the transfer learning process and preventing negative transfer. Extensive simulation studies and an application to predict ESRD demonstrate that the proposed method outperforms benchmarks such as target-only, source-only, and stratified Cox models, as well as other suitable transfer learning comparisons in the same setting.

The proposed two-step procedure in CoxTL is designed to ensure robustness against varying levels of data heterogeneity and potential model misspecification. This framework is highly adaptable, with multiple components that can be easily generalized. Specifically, the Cox model can be replaced with alternative time-to-event models, whether parametric or semiparametric, to suit different analytical needs. In the density ratio reweighting step, the exponential tilt model can be substituted with other statistical or machine learning approaches capable of estimating the ratio of two densities (Efron, 1978; Hastie, 2017; Sugiyama et al., 2012; Choi et al., 2021). A key strength of this approach lies in its ability to mitigate the impact of model misspecification by introducing a tuning parameter in the subsequent step. This parameter governs the influence of source data, effectively managing any adverse effects caused by mismatches in the earlier step.

The proposed CoxTL method, originally designed for a single-source setting, was extended to a multi-source framework in the real data application. In this extension, we applied density ratio reweighting separately for each source population and incorporated their weighted contributions into a unified log-likelihood function (See Section 1 ‘Multi-Source Framework’ of Supplementary Material). This multi-source CoxTL approach enabled us to leverage information from both Black and White populations simultaneously to improve survival prediction for the target Hispanic cohort and shows robust performance compared to the single-source CoxTL. While it is possible to assign a unique tuning parameter for each source, this can become computationally intensive as the number of sources grows. Alternative approaches to manage multi-source settings include ensemble learning and model aggregation, where CoxTL is applied to each source and the final risk score is a weighted combination of all single-source estimators. Weights can be determined using a validation dataset (Gu et al., 2022).

Our approach currently requires patient-level data from the source cohort, which may pose practical challenges due to data-sharing restrictions. Extending this method to a federated or distributed version could mitigate these concerns, allowing the model to operate across separate data sources without direct data sharing. Existing research on distributed Cox models, such as Li et al. (2022) and Bayle et al. (2023), provides a foundation for this extension, demonstrating the feasibility of transferring model parameters instead of data, which aligns well with privacy-preserving regulations and reduces the need for centralized data repositories.

## CONCLUSION

Our study demonstrates that the proposed CoxTL is a robust and effective transfer learning method for survival analysis, particularly in scenarios involving covariate shifts and heterogeneous regression parameters between source and target cohorts. Through extensive simulations and real-world applications, CoxTL consistently outperformed existing methods in both predictive accuracy and calibration, as evidenced by superior C-index and IBS values. Its ability to adjust for covariate and hazard function differences makes it highly relevant for real-world applications where source and target populations often exhibit significant variability. Its ability to adjust for multi-level data heterogeneity makes it a valuable tool for integrative survival analysis, especially in settings with limited data availability.

## ACKNOWLEDGMENT

This study is supported by NIH Grants R01GM148494, R01MH137218, and R01CA296289.

## DATA AVAILABILITY STATEMENT

The All of Us data used in this study can be registered for access through the All of Us Research Hub (https://www.researchallofus.org/).

## SUPPLEMENTARY MATERIALS

The supplementary file contains two parts. The first part elaborates on the joint calibrated log-likelihood function for multi-source CoxTL, with a pseudo algorithm for hyperparameter selection. The second one shows the covariate contribution in real data analysis using SHAP value. R-package for CoxTL implementation is publicly available from https://github.com/YUYING-LU/CoxTL with a detailed tutorial file.

