## Supplementary pdf for "Adaptive Transfer Learning for Time-to-Event Modeling with Applications in Disease Risk Assessment"

#### 1 MULTI-SOURCE FRAMEWORK

Our Cox_TL method can be extended to a multi-source framework. Assume we have $m \geq 2$ source cohorts $\mathcal{S}_{(1)}, \cdots, \mathcal{S}_{(M)}$, and one target cohort $\mathcal{T}$. The $m$-th source cohort has $N^{\mathcal{S}_{(m)}}$ independent observations, which is much larger than that of the target cohort $N^{\mathcal{T}}$, i.e. $N^{\mathcal{S}_{(m)}} \gg N^{\mathcal{T}}$. The other notations are similar to what we used in Section 2 (Method) in our paper.

**Step 1: Density ratio estimation for each source cohort**

For $m \in \{1, \cdots, M\}$, we calculate the density ratio between the $i$-th source cohort and the unique target cohort, i.e. $\omega_{(m)}(\mathbf{X}) = P^{\mathcal{T}}(\mathbf{X})/P^{\mathcal{S}_{(m)}}(\mathbf{X})$, where $P^{\mathcal{T}}(\cdot)$ and $P^{\mathcal{S}_{(m)}}(\cdot)$ denote the probability density function of $\mathbf{X}$ in the target cohort $\mathcal{T}$ and the $m$-th source cohort $\mathcal{S}_{(m)}$ respectively. We assume the density ratio model $\omega_{(m)}(\mathbf{X})$ follows the exponential tilting model with an underlying $p+1$ dimensional parameter $\boldsymbol{\theta}_{(m)} = (\theta_{(m),0}, \theta_{(m),1}, \cdots, \theta_{(m),p})^\top$:

$$\omega_{(m)}(\mathbf{X}) = \exp\{\mathbf{Z}^\top \boldsymbol{\theta}_{(m)}\},$$

where $\mathbf{Z} = (1, \mathbf{X})$ and we estimate $\boldsymbol{\theta}_{(m)}$ through

$$\hat{\boldsymbol{\theta}}_{(m)} = \arg\min_{\boldsymbol{\theta}} \Big\{ \frac{1}{N_{\mathcal{T}}} \sum_{i=1}^{N_{\mathcal{T}}} (\mathbf{Z}_i^{\mathcal{T}})^\top \boldsymbol{\theta} - \frac{1}{N_{\mathcal{S}_{(m)}}} \sum_{i=1}^{N_{\mathcal{S}_{(m)}}} \exp\{(\mathbf{Z}_i^{\mathcal{S}_{(m)}})^\top \boldsymbol{\theta}\} + \lambda_{(m)} \mathcal{P}_{\boldsymbol{\theta}_{(m)}} \Big\}.$$

$\mathcal{P}_{\boldsymbol{\theta}_{(i)}}$ can be any penalty function encouraging certain structures on $\boldsymbol{\theta}_{(m)}$, and $\lambda_{(m)}$ is the tuning parameter. In this study, we use L2 regularization as the penalty, i.e., $\mathcal{P}_{\boldsymbol{\theta}_{(m)}} = \sum_{i=0}^{p} \theta_{(m),i}^2$ and set $\lambda_{(m)} = 1$ as the default value for convenience.

**Step 2: Joint calibrated log-likelihood function**

Given the density ratio model, $\hat{\omega}_{(m)}(\mathbf{X}) = \exp\{\mathbf{Z}^\top \hat{\boldsymbol{\theta}}_{(m)}\}$, estimated in Step 1, we denote the re-weighted log-likelihood function in the $m$-th source cohort by

$$l_{\text{re}}^{\mathcal{S}_{(m)}}(\boldsymbol{\beta}) = \sum_{i \in \mathcal{D}^{\mathcal{S}_{(m)}}} \hat{\omega}_{(m)}(\mathbf{X}_i^{\mathcal{S}_{(m)}}) \big[ (\mathbf{X}_i^{\mathcal{S}_{(m)}})^\top \boldsymbol{\beta} + log\{\hat{h}_0^{\mathcal{S}_{(m)}}(Y_i^{\mathcal{S}_{(m)}} | \boldsymbol{\beta})\} \big],$$

where $\mathcal{D}^{\mathcal{S}_{(m)}} = \{i : \delta_i^{\mathcal{S}_{(m)}} = 1\}, m = 1, \cdots, M$, and

$$\hat{h}_0^{\mathcal{S}_{(m)}}(Y_i^{\mathcal{S}_{(m)}} | \boldsymbol{\beta}) = \frac{\omega_{(m)}(\mathbf{X}_i^{\mathcal{S}_{(m)}}) \cdot \delta_i^{\mathcal{S}_{(m)}}}{\sum_{j: Y_j^{\mathcal{S}_{(m)}} \geq Y_i^{\mathcal{S}_{(m)}}} \omega_{(m)}(\mathbf{X}_j^{\mathcal{S}_{(m)}}) \cdot exp\{(\mathbf{X}_j^{\mathcal{S}_{(m)}})^\top \boldsymbol{\beta}\}}, i \in \mathcal{D}^{\mathcal{S}_{(m)}}$$

is the calibrated baseline hazard estimation for the $m$-th source cohort. We proposed a joint calibrated log-likelihood function for multi-source setting:

$$l^{\text{TL\_joint}}(\boldsymbol{\beta}) = \underbrace{\sum_{i \in \mathcal{D}^{\mathcal{T}}} \left[ (\mathbf{X}_i^{\mathcal{T}})^\top \boldsymbol{\beta} + log\{\hat{h}_0^{\mathcal{T}}(Y_i^{\mathcal{T}}|\boldsymbol{\beta})\} \right]}_{\text{Target}} + \underbrace{\sum_{m=1}^{M} \nu_{(m)} \cdot l_{\text{re}}^{\mathcal{S}_{(m)}}(\boldsymbol{\beta})}_{\text{Jointly Weighted Source}},$$

The scalar hyperparameter $\nu_{(m)}$ is designed to control the degree of information sharing from the $m$-th source cohort to the target cohort, thereby preventing negative transfer learning caused by significant heterogeneity or potential model misspecifications. The values of $\{\nu_{(m)}\}_{m=1}^{M}$ are selected one-by-one via k-fold cross-validation on target cohort. The detailed process of choosing the hyperparameters via cross-validation is shown in Algorithm 1.

---

**Algorithm 1:** Cross-validation for selecting $\nu_{(m)}, m = 1, \cdots, M$

---

**Input:** $\{Y_i^{\mathcal{T}}, \delta_i^{\mathcal{T}}, \mathbf{X}_i^{\mathcal{T}}\}_{i=1}^{N^{\mathcal{T}}}$ and $\{Y_i^{\mathcal{S}_{(m)}}, \delta_i^{\mathcal{S}_{(m)}}, \mathbf{X}_i^{\mathcal{S}_{(m)}}\}_{i=1}^{N^{\mathcal{S}_{(m)}}}, m = 1, \cdots, M$.

1. Set initialization

**for** $m = 1, \cdots, M$ **do**

With single source log-likelihood function

$$l_{(m)}(\boldsymbol{\beta}) = \underbrace{\sum_{i \in \mathcal{D}^{\mathcal{T}}} \left[ (\mathbf{X}_i^{\mathcal{T}})^\top \boldsymbol{\beta} + log\{\hat{h}_0^{\mathcal{T}}(Y_i^{\mathcal{T}}|\boldsymbol{\beta})\} \right]}_{\text{Target}} + \underbrace{\nu_{(m)}^{\text{ini}} \cdot l_{\text{re}}^{\mathcal{S}_{(m)}}(\boldsymbol{\beta})}_{\text{Single Weighted Source}},$$

choose the hyperparameter $\nu_{(m)}^{\text{ini}}$ among grid $\mathcal{V}$ that gives the highest mean value of C-index via k-fold cross-validation and use that as the initial value for $\nu_{(k)}$.

**end**

2. Select $\nu_{(m)}$ one-by-one

**for** $m = 1, \cdots, M$ **do**

With joint log-likelihood

$$l_{(m)}^{\text{TL\_joint}}(\boldsymbol{\beta}) = \underbrace{\sum_{i \in \mathcal{D}^{\mathcal{T}}} \left[ (\mathbf{X}_i^{\mathcal{T}})^\top \boldsymbol{\beta} + log\{\hat{h}_0^{\mathcal{T}}(Y_i^{\mathcal{T}}|\boldsymbol{\beta})\} \right]}_{\text{Target}}$$
$$+ \underbrace{\sum_{m'<m} \nu_{(m')} \cdot l_{\text{re}}^{\mathcal{S}_{(m')}}(\boldsymbol{\beta}) + \nu_{(m)} \cdot l_{\text{re}}^{\mathcal{S}_{(m)}}(\boldsymbol{\beta}) + \sum_{m''>m} \nu_{(m'')}^{\text{ini}} \cdot l_{\text{re}}^{\mathcal{S}_{(m'')}}(\boldsymbol{\beta})}_{\text{Jointly Weighted Source}},$$

choose the hyperparameter $\nu_{(m)}$ among grid $\mathcal{V}$ that gives the highest mean value of C-index via k-fold cross-validation and use that as the final value for $\nu_{(m)}$.

**end**

**Output:** $\nu_{(m)}, m = 1, \cdots, M$

---

### 2 COVARIATE CONTRIBUTIONS

This section focuses on analyzing the contribution of each covariate (listed in Section 3 Table 1) that we use in real-data analysis. We apply SHAP analysis with respect to the following three groups – 'Target Hispanic', 'Source White', and 'Source Black', to separately evaluate the contribution of each covariate to ESRD risk prediction model in different populations. For each group, we select all the diseased cases and train an XGBoost model (with learning rate = 0.1 and boosting iterations =65) that aims at predicting the death time of each observation. The SHAP value of each covariate is available and we list the top 8 covariates with the highest SHAP values for each population in Figure 1.

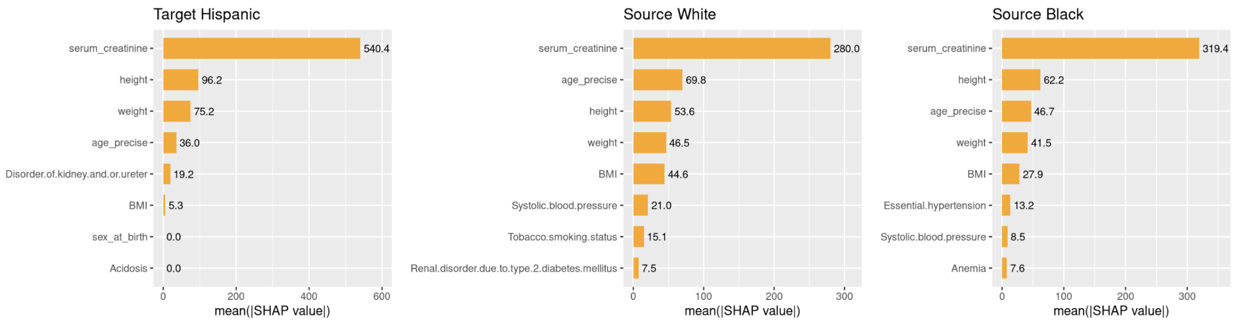


**Figure 1: The top 8 covariates contributing to the model's predictions based on SHAP values.**

The covariates 'serum_creatinine', 'height', 'weight', 'age_precise', and 'BMI' are consistently among the top eight covariates with the highest SHAP values across all three models, illustrating their significant contribution to ESRD prediction. The SHAP analysis identifies the most influential variables in the model, but it does not imply novelty in risk prediction. The high importance of serum creatinine is expected, given its central role in kidney function assessment. Similarly, BMI is correlated with height and weight; however, it is included as an established independent predictor of CKD progression. 'Diseases of the Kidney and Ureter' appears due to the hierarchical structure of ICD-10 codes, reflecting variations in disease severity rather than mere diagnosis presence. While acidosis is a modifiable state, its presence in historical EHR records serves as a marker of metabolic instability, which may contribute to ESRD progression.

### 3 COVARIATE DETAILS

In this section, we provide detailed information about the All of Us data used in our real-data analysis. Figure 2 presents the distribution of follow-up times for patients. Table 1 shows the names of all 60 covariates included in the ESRD risk prediction model.

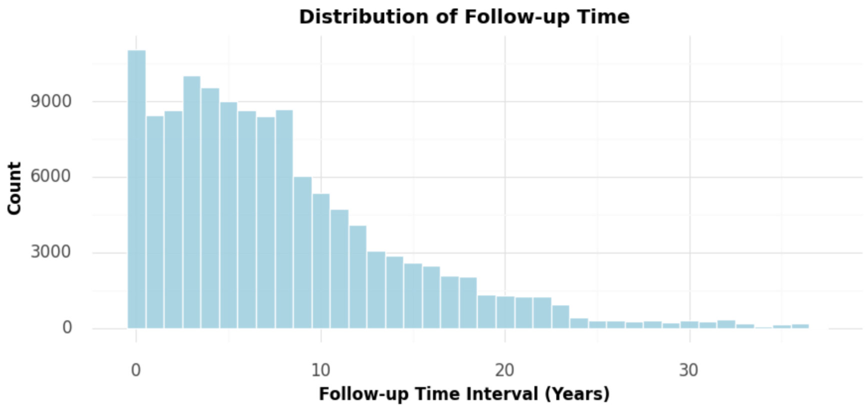


**Figure 2: Distribution of Follow-up Times of Patients.**

**Table 1: 60 Covariates in ESRD Risk Prediction Model.**

| Category | Variables |
| --- | --- |
| **Demographic Factors** | sex_at_birth, age_precise, height, weight, BMI |
| **Laboratory Measurements** | Systolic.blood.pressure, Low.blood.pressure, Hemoglobin.A1c.Hemoglobin.total.in.Blood, Creatinine.Mass.volume.in.Body.fluid, Iron.Mass.volume.in.Serum.or.Plasma, Iron.saturation.Mass.Fraction.in.Serum.or.Plasma, Iron.binding.capacity.Mass.volume.in.Serum.or.Plasma, Ferritin.Mass.volume.in.Serum.or.Plasma, Parathyrin.intact.Mass.volume.in.Serum.or.Plasma, Bilirubin.indirect.Mass.volume.in.Serum.or.Plasma, Erythrocyte.distribution.width.Ratio..by.Automated.count, Protein.Mass.volume.in.Urine, Hepatitis.C.virus.Ab.Presence.in.Serum.or.Plasma |
| **Kidney Function and Related Disorders** | serum_creatinine, Frank.hematuria, Proteinuria, Glomerular.filtration.rate, Nephritic.syndrome, Disorder.of.kidney.and.or.ureter, Acute.renal.failure.syndrome, Renal.disorder.due.to.type.1.diabetes.mellitus, Renal.disorder.due.to.type.2.diabetes.mellitus, Chronic.kidney.disease.stage.2, Chronic.kidney.disease.stage.3 |
| **Diabetes and Related Conditions** | Type.1.diabetes.mellitus, Type.2.diabetes.mellitus, Polyneuropathy.due.to.diabetes.mellitus, Hypoglycemia, insulin.glargine |
| **Cardiovascular and Vascular Disorders** | Atherosclerosis.of.coronary.artery.without.angina.pectoris, Congestive.heart.failure, Peripheral.vascular.disease, Peripheral.venous.insufficiency, Hypercoagulability.state Essential.hypertension |
| **Metabolic and Endocrine Disorders** | Disorder.of.phosphorus.metabolism, Disorder.of.parathyroid.gland, Hyperparathyroidism, Hypercalcemia, Hyperkalemia, Hypothyroidism, Gout |
| **Anemia and Hematologic Conditions** | Anemia, Anemia.in.chronic.kidney.disease, Anemia.of.chronic.disease, Iron.deficiency.anemia, Nutritional.anemia |
| **Other Conditions** | Acidosis, Altered.mental.status, Chronic.pain.syndrome, Sepsis, Systemic.lupus.erythematosus, highest_smoking_status_rank, Tobacco.smoking.status |
